# Discovery of a simple two-gene expression biomarker in whole blood predictive of the need for treatment escalation in inflammatory bowel disease

**DOI:** 10.1101/2021.07.09.21259804

**Authors:** Jan K. Nowak, Rahul Kalla, Alex T. Adams, IBD-Character Consortium, Jonas Halfvarson, Jack Satsangi

## Abstract

**Background and aims:** The IBD-Character consortium has recruited large internationally based inception cohorts of treatment-naïve inflammatory bowel disease (IBD) patients, providing a unique resource to derive a simple transcriptome signature in the field of prognostication.

**Methods:** The discovery cohort (n=160) was recruited in Norway, Sweden and Spain. The replication inception cohort from the United Kingdom (n=97) was followed-up for a mean (±SD) of 350±228 days. Treatment escalation was formally defined as the need for a biologic agent, ciclosporin and/or surgery, instituted for disease flare after initial remission, or colectomy during the index admission for ulcerative colitis. Whole blood RNA was subject to paired-end sequencing. In the discovery cohort a simple procedure was applied, which exploited differences of transcript ratios. The ten top performing ratios were tested using Cox regression models in the validation cohort.

**Results:** Newly diagnosed IBD patients with high *CACNA1E/LRRC42* expression ratio had an increased risk of treatment intensification (validation cohort: HR=19.3, 95%CI 2.6–143.9, p=0.000005; AUC 0.76, 95%CI 0.66–0.86). In 51 patients with CRP < 3.5 mg/L, *CACNA1E/LRRC42* still predicted escalation (HR=10.4; 95%CI 1.2-86.5, p=0.007). The second best performing transcript ratio was *CACNA1E/CEACAM21* yielding a HR of 10.9 (95%CI 2.5-46.7, p=0.00002) and an AUC of 0.76 (95%CI 0.65-0.86) in the validation cohort.

**Conclusion:** Transcriptomic profiling of an IBD inception cohort identified gene expression ratios *CACNA1E/LRRC42* and *CACNA1E/CEACAM21* as prognostic biomarkers. These were validated in a replication cohort as strongly associated with short- and long-term risk of treatment intensification and may provide valuable information in clinical decision-making.

## Introduction

The field of therapeutics in inflammatory bowel disease (IBD) has seen marked expansion over the last decade. In parallel, expectations have developed that biomarker discovery may help in rationalizing therapeutic choice in individual patients. A number of multi-omic analyses have been proposed to be of potential clinical use in predicting disease course in IBD. Discoveries in recent studies include genetic, epigenetic, transcriptomic and proteomic signals associated with treatment escalation at diagnosis^1,2^. Of particular note is the whole blood-derived transcriptomic signature, described by Biasci and colleagues, which is now being assessed in a prospective clinical trial in Crohn’s disease^3^ (CD).

The IBD-Character consortium has recruited large internationally based inception cohorts of treatment-naïve IBD patients with extensive multi-omic characterization, thereby providing a unique resource to derive a simple transcriptome signature in the field of prognostication. In this context, IBD-Character has to date enabled the identification of prognostic proteomic and micro-RNA biomarkers^2,4^. In these cohorts, we now show that simple gene expression signatures in whole blood taken at initial presentation can define future disease course and provide complementing data to be used in clinical practice.

## Methods

The discovery cohort (n=160) was recruited in Oslo (Norway; n=87), Örebro (Sweden; n=52), and Zaragoza (Spain; n=21) and included newly diagnosed patients who escalated treatment within the first year of diagnosis or did not require escalation but were followed-up for at least a year (Supplementary Table 1).

In the replication inception cohort from Edinburgh (United Kingdom; n=97) the mean (± SD) follow-up was 350 ± 228 days. All the centers employed the step-up treatment strategy. Treatment escalation was formally defined as the need for a biologic agent, ciclosporin and/or surgery, instituted for disease flare after initial remission. In ulcerative colitis (UC), the definition of treatment escalation also included any patient requiring colectomy during the index admission.

Whole blood was collected to Paxgene tubes, and the obtained RNA was subject to paired-end sequencing (AmpliSeq). In the discovery cohort, reads were filtered for highly expressed genes only (n=13,071), and a simple procedure was applied, which exploited differences of transcript ratios (described in detail in Supplementary Methods). In the validation cohort, the signals were tested in all 21,913 annotated transcripts using Cox regression models.

## Results

Eighty-five million ratios were analyzed to identify the top 10 ratios with the highest areas under the curve (AUCs) in the discovery cohort; of these, two ratios demonstrated highly significant validation in the replication cohort (Figure 1). Six other ratios were validated with smaller effects (Supplementary Methods).

**Figure 1.**
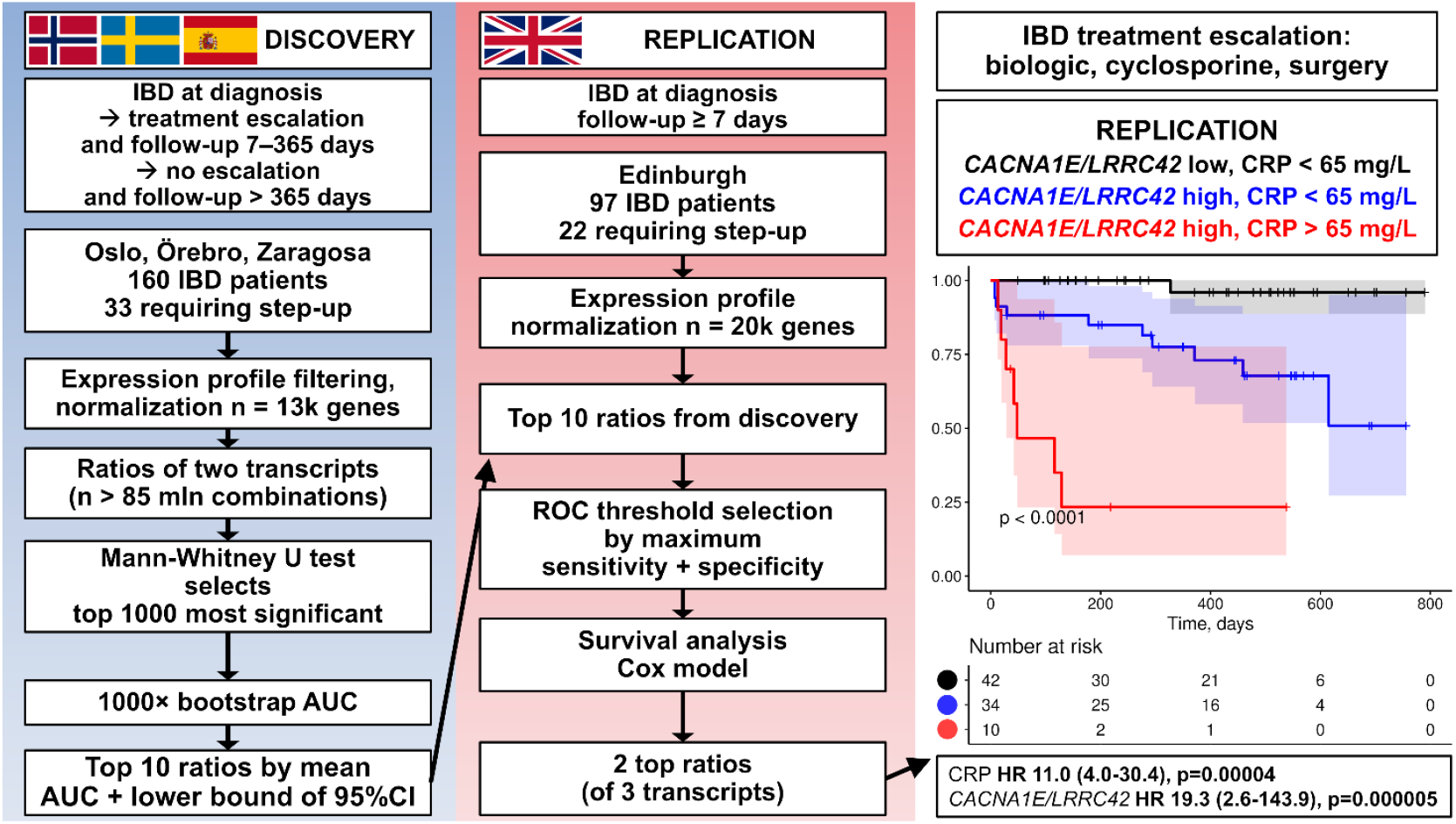
**(Left)** Overview of the discovery procedure involving comparison of transcript ratios along with area under the curve (AUC) bootstrap procedure. **(Middle)** Replication of top 10 ratios from the discovery step was attempted and most highly significant results were obtained for two ratios (*CACNA1E/LRRC42, CACNA1E/CEACAM21*). **(Right)** *CACNA1E/LRRC42* performs better than CRP in identification of newly diagnosed patients with IBD at risk of future treatment escalation. CRP cut-off 65 mg/L was selected automatically as maximizing the sum of sensitivity and specificity. The capacity of *CACNA1E/LRRC42* to identify at-risk patients among those with low-CRP persists even after the CRP threshold is set at 3.5 mg/L (see main text). Confidence intervals (95%) for the survival curves are shown.

Newly diagnosed IBD patients with high *CACNA1E/LRRC42* expression ratio (Calcium Voltage-Gated Channel Subunit Alpha1 E / Leucine Rich Repeat Containing 42; NM_001205293/NM_052940 > 0.066) had an increased risk of treatment intensification (validation cohort: HR 19.3, 95%CI 2.6–143.9, p=0.000005; AUC 0.76, 95%CI 0.66–0.86). Furthermore, the ratio was complementary to baseline C-reactive protein (CRP) in prediction. All patients with CRP above a cut-off based on maximum sensitivity and specificity (>65 mg/L) also had high *CACNA1E/LRRC42*. However, importantly among patients with lower CRP concentrations, *CACNA1E/LRRC42* was able to identify a validation subgroup predisposed to escalation in the long term (Figure 1). CRP alone (>65 mg/L) provided a HR of 11.0 (95%CI 4.0-30.4, p=0.00004) and AUC of 0.66 (95%CI 0.50-0.83). In 51 patients with CRP < 3.5 mg/L, *CACNA1E/LRRC42* still predicted escalation; HR 10.4 (95%CI 1.2-86.5, p=0.007; Supplementary Figure 1). Adding albumin did not improve the performance of *CACNA1E/LRRC42*-based model.

The second best performing transcript ratio was *CACNA1E/CEACAM21* (*CACNA1E* / Carcinoembryonic Antigen-Related Cell Adhesion Molecule 21; NM_001205293/NM_001098506 > 0.037) yielding a HR of 10.9 (95%CI 2.5-46.7, p=0.00002) and an AUC of 0.76 (95%CI 0.65-0.86) in the validation cohort.

After adjustment for CRP, drug-naïve status, age and sex, *CACNA1E/LRRC42* retained HR of 12.4 (95%CI 1.6–96.0; p=0.000004) and *CACNA1E/CEACAM21*-associated HR was 6.9 (95%CI 1.5–31.8, p=0.00001), both in the validation cohort.

## Discussion

In clinical practice, CRP has been shown to be the most accessible prognostic biomarker in blood with respect to future treatment escalation in patients with IBD^5^, whilst calprotectin is the most informative faecal biomarker. We confirm the predictive ability of CRP in this context. Additionally, we demonstrate that in patients in whom CRP is not elevated, an at-risk subgroup can be discerned using the gene expression ratio *CACNA1E/LRRC42*. Crucially, even patients who required treatment escalation after as long as two years were identified at diagnosis using *CACNA1E/LRRC42*.

This finding provides renewed optimism for the potential for clinical translation of transcriptomic IBD biomarkers in view of current concerns^6^ regarding the organizational and technological complexity of replication efforts, as well as their cost and the intricacy of models. Our findings set the stage for future validation of this simple, low-cost transcript ratio that could be easily translated to the clinic.

Mechanisms underlying the observed relationships remain to be characterized. In CD, a polymorphism in *CACNA1E* has been proposed to correlate with progression from inflammatory to stricturing or penetrating phenotype^7^. Further work is needed to elucidate its role in IBD. As the *CACAN1E* blocker topiramate was inefficacious in IBD, it appears that either *CACNA1E* plays no causative role or that a more refined approach is required to target its function specifically in leukocytes. *LRRC42* is expressed in a broad range of cells, including enterocytes, memory T cells, goblet and tuft cells (PanglaoDB). The presence of CUL3 and KAT5 among its interactors (BioGRID) hints at involvement in autophagy. Finally, CEACAM21 is found in lymphoid tissues (EMBL-EBI Expression Atlas) and is the only transmembrane CEA family molecule that does not possess a motif allowing tyrosine-based activation or inhibition. Its expression correlates with poor lung cancer survival, suggesting a role in promoting immune tolerance^8^.

In conclusion, transcriptomic profiling of an IBD inception cohort identified gene expression ratios *CACNA1E/LRRC42* and *CACNA1E/CEACAM21* as prognostic biomarkers. These were validated in a replication cohort as strongly associated with short- and long-term risk of treatment intensification and may provide valuable information in clinical decision-making.

## Data Availability

The data supporting the findings of this study will be available from the authors upon reasonable request.

## Author contributions

Study design: JKN, RK, ATA, JH, JS. Patient recruitment and sample processing: RK, ATA, JH, JS. Data analysis: JKN, RK, ATA, JH, JS. Drafting the manuscript: JKN, RK. Study coordination: JH, JS. All authors were involved in critical review, editing, revision and approval of the final manuscript.

---

**Supplementary Table 1.** Characteristics of the discovery and replication cohorts.

|  | DISCOVERY |  | REPLICATION |  |
| --- | --- | --- | --- | --- |
|  | Treatment escalation | No treatment escalation | Treatment escalation | No treatment escalation |
| n (% of cohort) | 33 (21%) | 127 (70%) | 22 (23%) | 75 (77%) |
| Oslo | 17 (52%) | 70 (55%) | - | - |
| Örebro | 10 (30%) | 42 (33%) | - | - |
| Zaragoza | 6 (18%) | 15 (12%) | - | - |
| Edinburgh | - | - | 22 (100%) | 75 (100%) |
| Follow-up, days, mean ± SD | 125 ± 81 | 809 ± 292 | 160 ± 170 | 406 ± 213 |
| Age, years, mean ± SD | 28.6 ± 10.7 | 36.6 ± 12.6 | 31.9 ± 11.5 | 37.1 ± 13.4 |
| Female | 16 (48%) | 64 (50%) | 6 (27%) | 32 (43%) |
| CD, n | 22 (67%) | 38 (30%) | 11 (50%) | 31 (41%) |
| Age at CD diagnosis | A2: 19 (86%)<br>A3: 3 (14%) | A2: 30 (81%)<br>A3: 7 (19%) | A2: 8 (73%)<br>A3: 3 (27%) | A2: 22 (71%)<br>A3: 9 (29%) |
| CD location | L1: 7 (32%)<br>L2: 5 (23%)<br>L3: 10 (45%)<br>L4: 0 | L1: 11 (29%)<br>L2: 12 (43%)<br>L3: 15 (39%)<br>L4: 22 (58%) | L1: 4 (36%)<br>L2: 3 (27%)<br>L3: 4 (36%)<br>L4: 0 (0%) | L1: 12 (39%)<br>L2: 9 (29%)<br>L3: 9 (29%)<br>L4: 1 (3%) |
| CD behavior | B1: 17 (77%)<br>B2: 4 (18%)<br>B3: 1 (5%) | B1: 35 (94%)<br>B2: 1 (3%)<br>B3: 1 (3%) | B1: 8 (73%)<br>B2: 1 (9%)<br>B3: 2 (18%) | B1: 17 (55%)<br>B2: 3 (10%)<br>B3: 4 (13%)<br>BB1: 6 (19%) |
| CD perianal | P: 1 (5%) | P: 1 (3%) | P: 1 (9%) | P: 6 (19%) |
| UC, n | 11 (33%) | 76 (60%) | 10 (45%) | 37 (49%) |
| UC extent | E2: 3 (27%)<br>E3: 8 (73%) | E1: 23 (30%)<br>E2: 26 (34%)<br>E3: 27 (36%) | E2: 2 (20%)<br>E3: 8 (80%) | E1: 13 (35%)<br>E2: 11 (30%)<br>E3: 13 (35%) |
| IBD-U | 0 | 13 (10%) | 1 (5%) | 7 (9%) |
| CRP, mg/L, mean ± SD | 51.7 ± 73.4 | 18.0 ± 38.2 | 58.2 ± 78.4 | 10.3 ± 20.5 |
| Albumin, g/L, mean ± SD | 34.0 ± 7.2 | 38.3 ± 5.9 | 34.2 ± 7.6 | 37.9 ± 5.6 |
| Smoking, n | All: 5 (15%)<br>CD: 5 (23%)<br>UC: 0 | All: 20 (16%)<br>CD: 13 (34%)<br>UC: 6 (8%) | All: 8 (36%)<br>CD: 5 (45%)<br>UC: 2 (20%) | All: 14 (19%)<br>CD: 11 (35%)<br>UC: 2 (5%) |
| Treatment-naïve, n (%) | 25 (76%) | 98 (77%) | 7 (32%) | 33 (44%) |

| Medication administered prior to inclusion (listed below) |  |  |  |  |
| --- | --- | --- | --- | --- |
| Oral 5-ASA | 1 (3%) | 1 (1%) | 5 (23%) | 12 (16%) |
| Rectal 5-ASA | 0 | 4 (3%) | 0 | 14 (19%) |
| Rectal steroids | 0 | 3 (2%) | 1 (4%) | 5 (7%) |
| Azathioprine / 6-MP | 0 | 1 (1%) | 0 | 2 (3%) |
| Methotrexate | 0 | 0 | 0 | 0 |
| Cyclosporine | 0 | 0 | 2 (9%) | 0 |
| Metronidazole | 0 | 1 (1%) | 3 (14%) | 6 (8%) |
| Ciprofloxacin | 1 (3%) | 2 (2%) | 1 (4%) | 3 (4%) |
| Other antibiotics | 1 (3%) | 5 (4%) | 9 (40%) | 26 (34%) |
| Intravenous steroids | 1 (3%) | 3 (2%) | 5 (23%) | 6 (8%) |
| High-dose prednisone | 3 (9%) | 3 (2%) | 4 (18%) | 2 (3%) |
| Low-dose prednisone | 0 | 0 | 1 (4%) | 2 (3%) |
| Infliximab | 1 (3%) | 1 (1%) | 0 | 0 |
| Adalimumab | 0 | 0 | 0 | 0 |

5-ASA – 5-aminosalicylates, 6-MP – 6-mercaptopurine, CD – Crohn's disease, CRP – C-reactive protein, IBD-U – inflammatory bowel disease unclassified, NSAID – nonsteroidal anti-inflammatory drug, TNF – tumor necrosis factor $\alpha$ , UC – ulcerative colitis

**Supplementary Figure 1.**
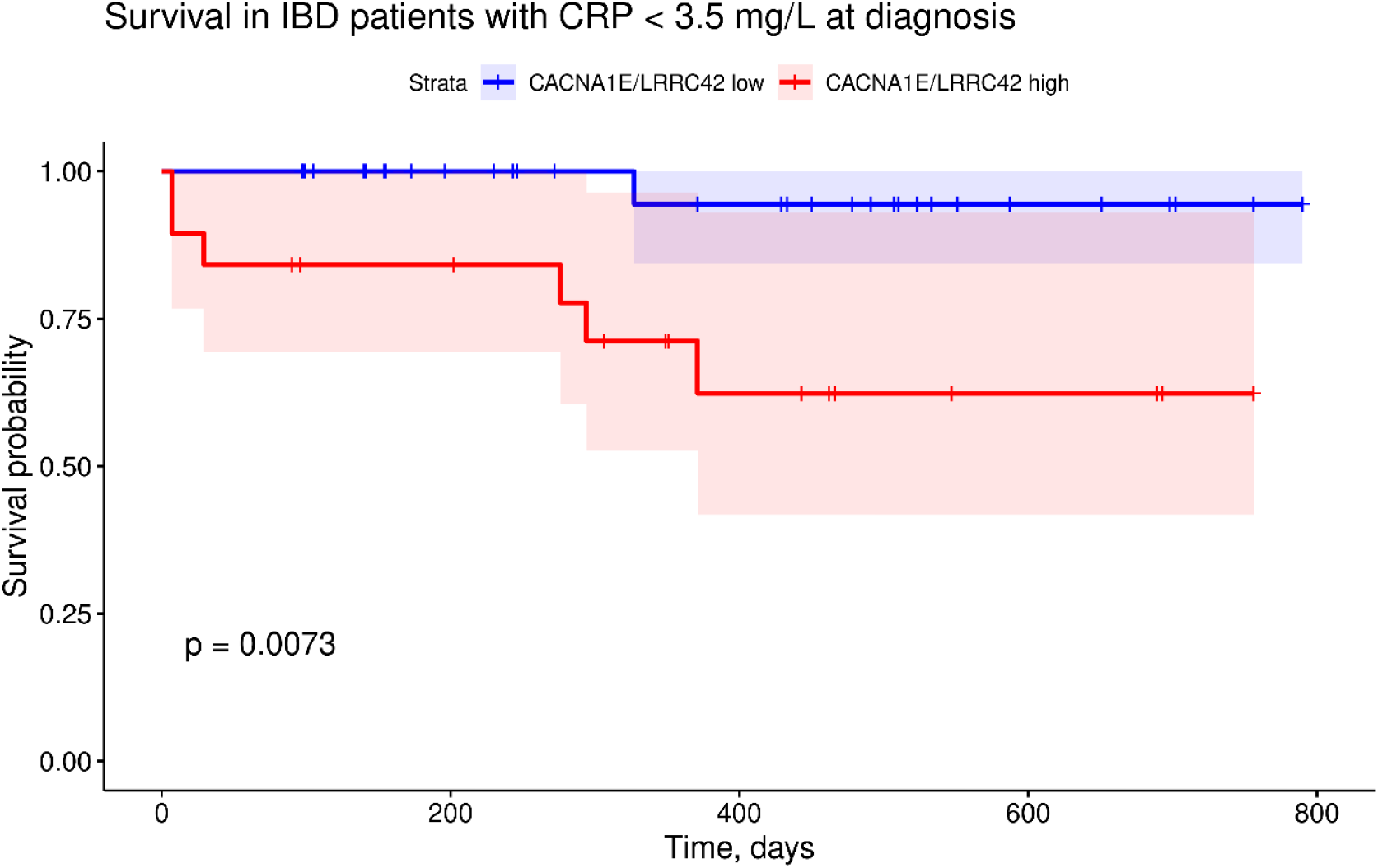
In the replication cohort, *CACNA1E/LRRC42* was able to identify patients at risk of treatment escalation among 51 persons with CRP < 3.5 mg/L (a highly-performing threshold that we determined previously in Kalla R et al. Am J Gastroenterol. 2016; 111:1796-1805). Low *CACNA1E/LRRC42* strongly associates with the absence of the need for escalation, even in the longer term (HR = 10.4; 95%CI 1.2-86.5, p=0.007).

## Supplementary methods

The patients were recruited in 2013-2016. All the participants gave informed written consent for their inclusion in the study. Whole blood was collected to Paxgene tubes. The obtained RNA was quantified (Qubit 2.0), checked for quality (RNA integrity number) and finally subject to paired-end sequencing using Ion AmpliSeq Human Gene Expression Core Panel (Thermo Fisher Scientific, Waltham, USA). Each pool contained eight equimolar samples. A library quality check was performed with the Ion Library TaqmanTM Quantitation kit (Thermo Fisher Scientific). Alignment of reads to hg19 was achieved with Torrent Software v. 4.6.

Four patients with mismatched sex were removed, as were: one patient with “possible IBD” diagnosis and one patient with “other inflammation” diagnosis. High-sensitivity CRP was assayed in one batch. Smoking status was self-declared. Analyses were conducted in R 4.0.4 (R Software Foundation, Vienna, Austria).

### Discovery cohort filtering and normalization

Patients from Oslo, Örebro and Zaragoza had a follow-up period of 7–365 days and treatment escalation or >365 days and no escalation detected at any timepoint. The data were checked for duplicates. Genes with raw mean expression ≥10 and less than 5% of null values were processed further. Globin genes were removed. After obtaining gene names using *org*.*Hs*.*eg*.*db* package, *biomaRt* was queried for gene lengths, enabling transcripts-per-million (TPM) normalization using code adapted from *bedapub/ribiosNGS*. The mean sample correlations were all >0.80.

### Discovery procedure

In the discovery cohort, all transcript ratios (∼85 mln) were compared between patients who escalated and who did not, using the fast implementation of two sample Wilcoxon test from the *matrixTests* package. Ratios including genes correlating with CRP (r>0.5) or each other (r>0.5) were excluded.

For the top 1000 ratios with the lowest p values, areas under the receiver operating characteristic curves (AUCs) were calculated using a 1000x bootstrap procedure, randomly selecting (with replacement) 10 escalating vs 10 non-escalating patients. AUC was calculated with the *pROC* package. The ratios were ranked by the sum of mean AUC and the 2.5^th^ centile. The top 10 ratios were selected for validation (*SPAG1/CACNA1E*\*\*\*, *CACNA1E/SPAG1*\*\*\*, *LOXL1/ BAIAP3*\*, *SPAG1/ FCGR1B*\*\*, *LOXL1/COL4A3, BAIAP3/LOXL1*\*, *COL4A3/LOXL1, CACNA1E/CEACAM21*\*\*\*, *LRRC42/FCGR1B*\*\*\*, *CACNA1E/LRRC42*\*\*\*; * - replication significance).

### Replication cohort normalization

Gene expression profiles of patients from Edinburgh with follow-up ≥7 days were included. Globin genes were removed, gene lengths were obtained as above, and the data was TPM-normalized as in *bedapub/ribiosNGS*. PCA and correlation plots were inspected. All the average sample correlations were >0.80.

### Validation in the replication cohort

In the replication cohort, the optimal cut-off was selected for each ratio by the criterion of maximum sum of sensitivity and specificity based on output from *pROC*. Packages *survival* and *survminer* were employed to analyze survival and prepare figures. Cox models were built, and from the total of ten ratios, the top two best performing models (lowest p) were selected. *CACNA1E/LRRC42* and CRP performance were visualized in the Edinburgh cohort. CRP threshold of 65 mg/L was automatically determined in the replication group by identifying the maximum sum of sensitivity and specificity. To complement these analyses, another threshold of 3.5 mg/L was used (which we determined as highly accurate in Kalla R et al. Am J Gastroenterol. 2016; 111:1796-1805).

## References

1. Biasci D, Lee JC, Noor NM, et al. A blood-based prognostic biomarker in IBD. Gut 2019;68:1386–1395.

2. Kalla R, Adams AT, Bergemalm D, et al. Serum proteomic profiling at diagnosis predicts clinical course, and need for intensification of treatment in inflammatory bowel disease. J Crohns Colitis 2021;15:699–708.

3. Parkes M, Noor NM, Dowling F, et al. PRedicting Outcomes For Crohn’s dIsease using a moLecular biomarkEr (PROFILE): protocol for a multicentre, randomised, biomarker-stratified trial. BMJ Open 2018;8:e026767.

4. Kalla R, Adams AT, Ventham NT, et al. Whole blood profiling of T-cell derived miRNA allows the development of prognostic models in inflammatory bowel disease. J Crohns Colitis 2020.

5. Kalla R, Kennedy NA, Ventham NT, et al. Serum Calprotectin: A Novel Diagnostic and Prognostic Marker in Inflammatory Bowel Diseases. Am J Gastroenterol 2016;111:1796–1805.

6. Gasparetto M, Payne F, Nayak K, et al. Transcription and DNA Methylation Patterns of Blood-Derived CD8+ T Cells Are Associated With Age and Inflammatory Bowel Disease But Do Not Predict Prognosis. Gastroenterology 2020.

7. Pernat Drobež C, Repnik K, Gorenjak M, et al. DNA polymorphisms predict time to progression from uncomplicated to complicated Crohn’s disease. Eur J Gastroenterol Hepatol 2018;30:447–455.

8. Nakamura H, Sakai H, Miyazawa T, et al. Differential prognostic values of mRNA expression of CEACAM gene family members in nonsmall cell lung cancer. Curr Biomark Find 2016;6:23–30.

